# Factors affecting COVID-19 outcomes in cancer patients: A first report from Guy’s Cancer Centre in London

**DOI:** 10.1101/2020.05.12.20094219

**Authors:** B Russell, C Moss, S Papa, S Irshad, P Ross, J Spicer, S Kordasti, D Crawley, H Wylie, F Cahill, A Haire, K Zaki, F Rahman, A Sita-Lumsden, D Josephs, D Enting, M Lei, S Ghosh, C Harrison, A Swampillai, E Sawyer, A Dsouza, S Gomberg, P Fields, D Wrench, K Raj, M Gleeson, K Bailey, R Dillon, M Streetley, A Rigg, R Sullivan, S Dolly, M Van Hemelrijck

## Abstract

**Background:** There is insufficient evidence to support clinical decision-making for cancer patients diagnosed with COVID-19 due to the lack of large studies.

**Methods:** We used data from a single large UK Cancer Centre to assess demographic/clinical characteristics of 156 cancer patients with a confirmed COVID-19 diagnosis between 29 February-12 May 2020. Logistic/Cox proportional hazards models were used to identify which demographic and/or clinical characteristics were associated with COVID-19 severity/death.

**Results:** 128 (82%) presented with mild/moderate COVID-19 and 28 (18%) with severe disease. Initial diagnosis of cancer >24m before COVID-19 (OR:1.74 (95%CI: 0.71-4.26)), presenting with fever (6.21 (1.76-21.99)), dyspnoea (2.60 (1.00-6.76)), gastro-intestinal symptoms (7.38 (2.71-20.16)), or higher levels of CRP (9.43 (0.73-121.12)) were linked with greater COVID-19 severity. During median follow-up of 47d, 34 patients had died of COVID-19 (22%). Asian ethnicity (3.73 (1.28-10.91), palliative treatment (5.74 (1.15-28.79), initial diagnosis of cancer >24m before (2.14 (1.04-4.44), dyspnoea (4.94 (1.99-12.25), and increased CRP levels (10.35 (1.05-52.21)) were positively associated with COVID-19 death. An inverse association was observed with increased levels of albumin (0.04 (0.01-0.04).

**Conclusions:** A longer-established diagnosis of cancer was associated with increasing severity of infection as well as COVID-19 death, possibly reflecting effects of more advanced malignant disease impact on this infection. Asian ethnicity and palliative treatment were also associated with COVID-19 death in cancer patients.

**Contribution to the field:** In the context of cancer, the COVID-19 pandemic has led to challenging decision-making. These are supported by limited evidence with small case studies being reported from China, Italy, New York and a recent consortium of 900 patients from over 85 hospitals in the USA, Canada, and Spain. As a result of their limited sample sizes, most studies were not able to distinguish between the effects of age, cancer, and other comorbidities on COVID-19 outcomes. Moreover, the case series from New York analysed which patient characteristics are associated with COVID-19 death, but only made a comparison with non-cancer patients. The first results of the COVID-19 and Cancer Consortium provide insights from a large cohort in terms of COVID-19 mortality, though a wide variety of institutions with different COVID-19 testing procedures were included.

Given the current lack of (inter)national guidance for cancer patients in the context of COVID-19, we believe that our large cancer centre can provide an important contribution to the urgent need for further insight into the intersection between COVID-19 and cancer. With comprehensive in-house patient details, consistent inclusion criteria and up-to-date cancer and COVID-19 outcomes, we are in position to provide rapid analytical information to the oncological community.

## Introduction

In the context of cancer, the COVID-19 pandemic has led to challenging decision-making (1, 2). Patient visits to the cancer clinic increase potential risk of infection when the alternative is self-isolation at home, and some cancer treatments may predispose patients to moderate or severe harmful effects of COVID-19 (3, 4). Current precautionary management decisions being made for cancer patients are based on assumptions supported by limited evidence, based on small case series from China and Italy (5-13) and larger series from New York (14, 15) and a recent consortium of 900 patients from over 85 hospitals in the USA, Canada, and Spain (16). As a result of their limited sample sizes, most studies were not able to distinguish between the effects of age, cancer, and other comorbidities on COVID-19 outcomes in this population (17, 18). Moreover, the case series from New York analysed which patient characteristics are associated with COVID-19 death, but only made a comparison with non-cancer patients (14, 15). The first results of the COVID- 19 and Cancer Consortium provide insights from a large cohort in terms of COVID- 19 mortality, though a wide variety of institutions with different COVID-19 testing procedures were included (16). In addition, recently published prognostic studies in COVID-19 positive patients have been judged to be at high risk of bias, mainly due to non-representative selection of control patients, exclusion of patients who had not experienced the event of interest by the end of the study, high risk of model overfitting, and limited information on model building strategies used (19).

It can be difficult to confidently diagnose COVID-19 symptoms in cancer patients, as presenting features of the infection are often similar to cancer symptoms and treatment-related adverse events (17, 20). This may result in a delayed or missed COVID-19 diagnosis, which could lead to confounding of case and infection mortality rates, as well as late interventions for more life-threatening disease (21). In addition, COVID-19 may be a barrier to dignified and humane end-of-life cancer care (17). Finally, the pandemic is causing huge service reconfiguration for both curative and palliative oncology care, resulting in fewer clinic visits due to social distancing (22), cessation of screening, and delays or changes in treatments that will inevitably have serious impacts on cancer-related mortality and morbidity (17, 21). Our recent systematic review reported there is currently no definitive evidence that specific cytotoxic drugs are contraindicated in cancer patients infected with COVID-19 (23).

Larger studies with multivariate models are urgently warranted to further explore this intersection of COVID-19 and cancer in terms of clinical outcomes, so as to inform oncological care during this outbreak and potential future pandemics (24). Guy’s Cancer Centre in South-East London, which treats approximately 8,800 patients annually, including 4,500 new diagnoses, is one of the largest Comprehensive Cancer Centres in the UK and is currently at the epicentre of the UK COVID-19 epidemic.

## Methods

### Study population

Guy’s Cancer Cohort, a research ethics committee approved research database (Reference number: 18/NW/0297) of all routinely collected clinical data of cancer patients at Guy’s and St Thomas’ NHS Foundation Trust (GSTT), forms the basis of this observational study (25). The database contains routinely collected prospective and retrospective demographic/clinical data on all cancer patients treated at Guy’s Cancer Centre. We have an established clinical database for all cancer patients tested for COVID-19 either in outpatient clinics or ward setting since 29 February 2020. Using the unique hospital number, these databases were merged prior to anonymization for research purposes. We assessed outcomes included in the core outcome sets currently being developed for COVID-19 to ensure all relevant information is collected in our COVID specific database (26).

We have included cancer patients who received a diagnosis of COVID-19, from a positive PCR test, from 29^th^ February-12^th^ May 2020. Until 30 April 2020, a COVID- 19 test was ordered for cancer patients if they presented with symptoms necessitating hospitalization or if they were scheduled to undergo a cancer-related treatment. From 1 May 2020, COVID-19 testing was introduced as part of standard care, with about 25% of patients being swabbed daily depending on staff and testing kit availability. A total of 1,507 patients were tested between 29 February and 12 May 2020, of whom 156 had COVID-19 (10%).

### Statistical methods

In this analysis of our data, we had three aims:

1. To describe demographic and clinical characteristics of COVID-19 positive cancer patients, in terms of their COVID-19 and cancer diagnoses.
2. To identify which demographic and/or clinical factors were associated with COVID-19 severity in cancer patients.
3. To identify which demographic and/or clinical factors were associated with COVID-19 death in cancer patients.

Descriptive statistics were used to address the first aim. Most variables had several categories for the purpose of these descriptive analyses, but were collapsed for the purpose of regression analyses due to the sample size of our cohort. Socioeconomic status (low, middle, high) was determined based on the English Indices of Multiple Deprivation for postcodes (27). Lymphocyte count (x10^9^) was categorized as ≤0.5, 0.6-0.8, 0.9-1.2, and >1.2 based on the Common Terminology Criteria for Adverse Events v.5 (CTCAE). For the other laboratory variables, we created tertiles instead of clinical cut-offs due to cancer patients already having abnormal values for most of these blood markers (Ferritin, C-reactive protein, and albumin). Radical treatment was defined as those patients with a chance of long-term survival or cure.

For the second aim, we conducted logistic regression analyses. Mild/moderate COVID-19 was defined as pneumonia with or without sepsis (i.e. those patients managed on the ward), whereas severe COVID-19 was defined as acute respiratory distress syndrome (ARDS) or septic shock (i.e. those patients where severity reaches criteria for Intensive Care Unit admission, if deemed clinically appropriate). These definitions were based on the WHO COVID-19 classification (28). The models used to quantify the association between each factor and COVID-19 severity were defined through a directed acyclic graph (DAG) (Figure 1 in Appendix). Each factor was individually set as the main exposure variable in the model in order to determine the minimal adjustments required for each factor (Table 1 in Appendix).

The third research aim was addressed with Cox proportional hazards regression analyses, whereby the models were defined as above (Table 1 in Appendix). Follow-up was defined from date of COVID testing until death or 12 May 2020.

All statistical analyses were conducted with STATA version 15.1

## Results

### Demographic and clinical characteristics of COVID-19 positive cancer patients

128 patients (82%) presented with mild/moderate COVID-19 and 28 patients (18%) with severe COVID-19 (Table 1). More patients were male (58%) and aged 60+ (68%; median age: 67). However, 14% of the cancer population was aged <50 years (n=21; median age: 41). When stratified by COVID grade, more male cancer patients presented with severe disease (68%). Most patients were from a lower socioeconomic background (81%). With respect to ethnicity, about half were White, 22% were Black (n=32) and 4% were of Asian (n=6) origin. When stratified by COVID grade, a slightly larger proportion of patients from a white ethnic background had severe COVID (57%). Hypertension was the most reported comorbidity (47%), followed by diabetes mellitus (22%), renal impairment (19%) and cardiovascular disease (19%). However, benign lung conditions were more commonly reported for those who presented with severe COVID-19 (29% vs 13% in those with mild COVID-19).

**Table 1:** Demographic characteristics of COVID-19 positive cancer patients.

|  | Total<br>(n=156) |  | WHO COVID Grade |  |  |  |
| --- | --- | --- | --- | --- | --- | --- |
|  |  |  | Mild/Moderate<br>(n=128) |  | Severe<br>(n=28) |  |
|  | n | % | n | % | n | % |
| **Sex** |  |  |  |  |  |  |
| Male | 90 | 57.70 | 71 | 55.50 | 19 | 67.90 |
| Female | 66 | 42.30 | 57 | 44.50 | 9 | 32.10 |
| **Age** |  |  |  |  |  |  |
| <50 | 21 | 13.50 | 16 | 12.50 | 5 | 17.90 |
| 50-59 | 29 | 18.60 | 24 | 18.80 | 5 | 17.90 |
| 60-69 | 43 | 27.60 | 35 | 27.30 | 8 | 28.60 |
| 70-79 | 35 | 22.40 | 28 | 21.90 | 7 | 25.00 |
| ≥80 | 28 | 17.90 | 25 | 19.50 | 3 | 10.70 |
| Mean (SD) | 65.18 (14.80) |  | 65.74 (14.39) |  | 62.62 (16.41) |  |
| **SES** |  |  |  |  |  |  |
| Low | 126 | 80.80 | 106 | 82.80 | 20 | 71.40 |
| Medium | 1 | 0.60 | 1 | 0.80 | 0 | 0.00 |
| High | 16 | 10.30 | 12 | 9.40 | 4 | 14 |
| Missing | 13 | 8.30 | 9 | 7.00 | 4 | 14.30 |
| **Ethnicity** |  |  |  |  |  |  |
| White British | 66 | 42.30 | 50 | 39.10 | 16 | 57.10 |
| White Other | 12 | 7.70 | 9 | 7.00 | 3 | 10.70 |
| Black Caribbean | 8 | 5.10 | 8 | 6.30 | 0 | 0.00 |
| Black African | 15 | 9.60 | 14 | 10.90 | 1 | 3.60 |
| Black Other | 12 | 7.70 | 9 | 7.00 | 3 | 10.70 |
| Asian | 6 | 3.80 | 4 | 3.10 | 2 | 7.10 |
| Mixed | 2 | 1.30 | 2 | 1.60 | 0 | 0.00 |
| Other | 5 | 3.20 | 4 | 3.10 | 1 | 3.60 |
| Unknown | 30 | 19.20 | 28 | 21.90 | 2 | 7.10 |
| **Comorbidities** |  |  |  |  |  |  |
| Hypertension | 74 | 47.40 | 63 | 49.20 | 11 | 39.30 |
| Diabetes Mellitus | 35 | 22.40 | 31 | 24.20 | 4 | 14.30 |
| Lung Conditions | 25 | 16.00 | 17 | 13.30 | 8 | 28.60 |
| Renal Impairment | 30 | 19.20 | 26 | 20.30 | 4 | 14.30 |
| Liver Conditions | 3 | 1.90 | 3 | 2.30 | 0 | 0.00 |
| CVD | 29 | 18.60 | 24 | 18.80 | 5 | 17.90 |
| Frailty | 10 | 6.40 | 6 | 4.70 | 4 | 14.30 |
| Chronic Steroid Use | 4 | 2.60 | 4 | 3.10 | 0 | 0.00 |
| **No. of Comorbidities** |  |  |  |  |  |  |
| 0 | 43 | 27.60 | 34 | 26.60 | 9 | 32.10 |
| 1 | 48 | 30.80 | 40 | 31.30 | 8 | 28.60 |
| 2 | 33 | 21.20 | 28 | 21.90 | 5 | 17.90 |
| 3+ | 32 | 20.50 | 26 | 20.30 | 6 | 21.40 |
| **Smoking history** |  |  |  |  |  |  |
| Never | 59 | 37.80 | 51 | 39.80 | 8 | 28.60 |
| Current | 11 | 7.10 | 9 | 7.00 | 2 | 7.10 |
| Ex-smoker | 39 | 25.00 | 31 | 24.20 | 8 | 28.60 |
| Unknown | 47 | 30.10 | 37 | 28.90 | 10 | 35.70 |
| **Medications** |  |  |  |  |  |  |
| Polypharmacy | 68 | 43.60 | 57 | 44.50 | 11 | 39.30 |
| NSAIDs | 20 | 12.80 | 16 | 12.50 | 4 | 14.30 |
| ACE/ARB | 33 | 21.20 | 29 | 22.70 | 4 | 14.30 |
| Beta-blockers | 24 | 15.40 | 20 | 15.60 | 4 | 14.30 |

The most frequently reported tumour types were urological/gynaecological (29%), followed by haematological (18%) and breast (15%) (Table 2). The first group (n=45) comprised 21 prostate, 8 renal, 5 bladder and 11 gynaecological cancers. Of the 28 haematological malignancies, 4 (14.3%) were myeloid and 24 (85.7%) were lymphoid. Of all cancer patients tested for COVID-19, 80 were positive after their cancer-related hospital admission (51%), of which 61 were solid tumours (76%) and 19 were haematological cancers (24%). When stratified by COVID-19 severity, the largest proportion of cancers presenting with severe COVID were haematological (36%). A large proportion of patients had advanced cancer (40% stage IV) and were diagnosed with their malignancy in the last 12 months (46%).

**Table 2:** Tumour characteristics of COVID-19 positive cancer patients.

|  | Total<br>(n=156) |  | WHO COVID Grade |  |  |  |
| --- | --- | --- | --- | --- | --- | --- |
|  |  |  | Mild/Moderate<br>(n=128) |  | Severe<br>(n=28) |  |
|  | n | % | n | % | n | % |
| **Cancer type** |  |  |  |  |  |  |
| Urological/Gynae | 45 | 28.80 | 39 | 30.50 | 6 | 21.40 |
| Gastro-intestinal | 21 | 13.50 | 19 | 14.80 | 2 | 7.10 |
| Haematological | 28 | 17.90 | 18 | 14.10 | 10 | 35.70 |
| Skin/Head & neck/Sarcoma | 10 | 6.40 | 9 | 7.00 | 1 | 3.60 |
| Central Nervous System | 11 | 7.10 | 10 | 7.80 | 1 | 3.60 |
| Breast | 24 | 15.40 | 21 | 16.40 | 3 | 10.70 |
| Lung | 17 | 10.90 | 12 | 9.40 | 5 | 17.90 |
| **Cancer stage** |  |  |  |  |  |  |
| I | 17 | 10.90 | 17 | 13.30 | 0 | 0.00 |
| II | 23 | 14.70 | 21 | 16.40 | 2 | 7.10 |
| III | 22 | 14.10 | 18 | 14.10 | 4 | 14.30 |
| IV | 63 | 40.40 | 49 | 38.30 | 14 | 50.00 |
| Missing | 31 | 19.90 | 23 | 18.00 | 8 | 28.60 |
| **Risk category\* (n=4)** |  |  |  |  |  |  |
| Low | 1 | 25.00 | 0 | 0.00 | 1 | 33.33 |
| Intermediate | 2 | 50.00 | 1 | 100.00 | 1 | 33.33 |
| High | 1 | 25.00 | 0 | 0.00 | 1 | 33.33 |
| **Treatment Paradigm** |  |  |  |  |  |  |
| Treatment naive | 18 | 11.50 | 16 | 12.50 | 2 | 7.10 |
| Neoadjuvant | 7 | 4.50 | 7 | 5.50 | 0 | 0.00 |
| Adjuvant | 8 | 5.10 | 8 | 6.30 | 0 | 0.00 |
| Radical | 38 | 24.40 | 28 | 21.90 | 10 | 35.70 |
| Palliative | 60 | 38.50 | 49 | 38.30 | 11 | 39.30 |
| Watch and wait | 7 | 4.50 | 7 | 5.50 | 0 | 0.00 |
| Surveillance | 12 | 7.70 | 10 | 7.80 | 2 | 7.10 |
| Missing | 6 | 3.80 | 3 | 2.30 | 3 | 10.70 |
| **Line of Palliative Treatment (N=54)** |  |  |  |  |  |  |
| 1 | 27 | 50.00 | 23 | 52.27 | 4 | 40.00 |
| 2 | 18 | 33.33 | 13 | 29.55 | 5 | 50.00 |
| 3 | 6 | 11.11 | 5 | 11.36 | 1 | 10.00 |
| 4 | 1 | 1.90 | 1 | 2.27 | 0 | 0.00 |
| Missing | 2 | 3.70 | 2 | 4.55 | 0 | 0.00 |
| **Systemic Treatment (N=81)** |  |  |  |  |  |  |
| Systemic chemotherapy | 45 | 55.60 | 34 | 53.10 | 11 | 64.60 |
| Immunotherapy | 7 | 8.60 | 5 | 7.80 | 2 | 11.80 |
| Biological | 13 | 16.00 | 11 | 17.20 | 2 | 11.80 |
| Targeted Therapy | 5 | 6.20 | 5 | 7.80 | 0 | 0.00 |
| Combination Therapy | 11 | 13.60 | 9 | 14.10 | 2 | 11.80 |
| **Time since cancer diagnosis** |  |  |  |  |  |  |
| <3 months | 41 | 26.30 | 34 | 26.60 | 7 | 25.00 |
| 3-12 months | 30 | 19.20 | 25 | 19.50 | 5 | 17.90 |
| 12-24 months | 20 | 12.80 | 18 | 14.10 | 2 | 7.10 |
| >24 months | 57 | 36.50 | 45 | 35.20 | 12 | 42.90 |
| **Performance status** |  |  |  |  |  |  |
| 0 | 19 | 12.20 | 17 | 13.30 | 2 | 7.10 |
| 1 | 43 | 27.60 | 34 | 26.60 | 9 | 32.10 |
| 2 | 33 | 21.20 | 27 | 21.10 | 6 | 21.40 |
| 3 | 14 | 9.00 | 13 | 10.20 | 1 | 3.60 |
| 4 | 6 | 3.80 | 5 | 3.90 | 1 | 3.60 |

\*For myeloid malignancies only.

Overall, 39% of patients were receiving palliative treatment, 25% were receiving radical treatment and 12% were treatment naive. Treatment distributions were reasonably comparable between COVID-19 severity groups. Of the 81 patients on systemic treatment within the last 2 years, 54 were in a palliative setting, of these 50% were 1^st^ line, 33% 2^nd^ line and 13% on ≥3^rd^ treatment line. However, the majority of severe COVID-19 patients were on third line metastatic treatment. Table 2 provides further details on the cancer characteristics.

46% of the cancer patients diagnosed with COVID-19 in this cohort presented with a cough and 52% had a fever. Most patients were molecularly diagnosed within 7 days of their initial symptoms (58%) (Table 3). More patients in the severe COVID-19 group presented with C-reactive protein (CRP) values in the highest tertile (46 vs 22% for mild/moderate disease). Similarly, they had a lower lymphocyte count (53 vs 21% in the lowest category (≤0.5)) and lower albumin levels (39 vs 22% in the lowest tertile).

**Table 3:**
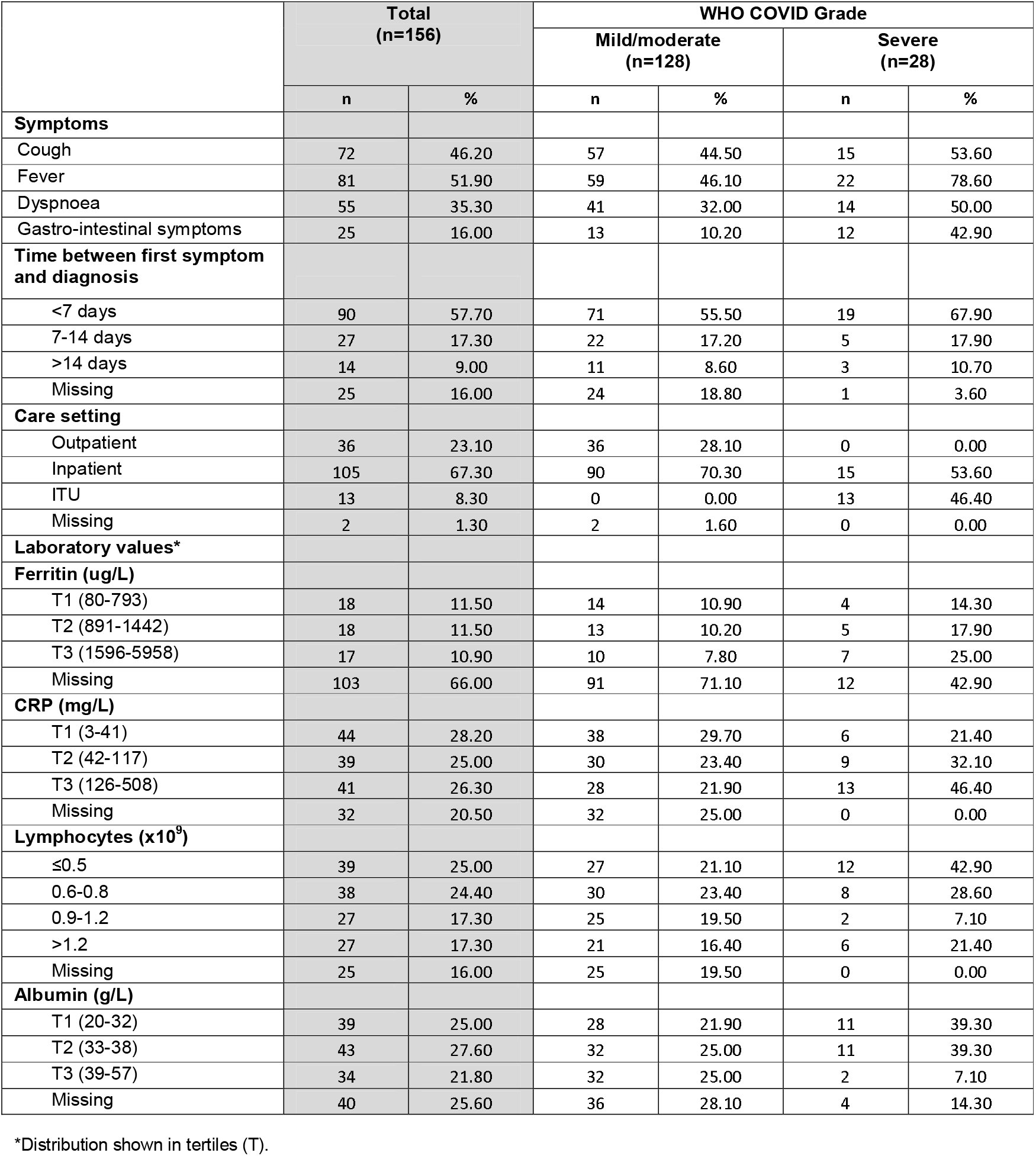
COVID-19 presentation of COVID-19 positive cancer patients.

### Factors associated with COVID-19 severity in cancer patients

The odds ratios (ORs) for the associations between the various demographic and clinical factors and COVID-19 severity status are shown in Table 4. There was a non-statistically significant indication that those patients who were diagnosed with cancer more than 24 months ago were at a higher risk of presenting with severe COVID-19 as compared to those diagnosed during the last 24 months (OR: 1.74 (95%CI:0.71-4.26)). With respect to symptom presentation, those presenting with a fever, dyspnoea, or gastro-intestinal symptoms were at a higher risk of having severe COVID-19 as compared to those without these symptoms (OR: 6.21 (1.76-21.99), 2.60 (1.00-6.76), and 7.38 (2.71-20.16), respectively).

**Table 4:** Odds Ratios and 95% Confidence intervals for COVID-19 severity in cancer patients.

|  | OR* | 95% CI |
| --- | --- | --- |
| **Sex** |  |  |
| Male | 1.00 | Ref |
| Female | 0.59 | (0.25-1.40) |
| **Age** |  |  |
| ≤60 | 1.00 | Ref |
| >60 | 0.82 | (0.35-1.93) |
| **SES** |  |  |
| Low | 1.00 | Ref |
| Middle | N/A | N/A |
| High | 2.11 | (0.51-8.67) |
| **Ethnicity** |  |  |
| White | 1.00 | Ref |
| Black | 0.40 | (0.13-1.28) |
| Asian | 1.55 | (0.26-9.16) |
| Other | 0.52 | (0.06-4.57) |
| **Number of comorbidities** |  |  |
| 0 | 1.00 | Ref |
| 1 | 1.09 | (0.29-4.07) |
| 2 | 1.04 | (0.23-4.68) |
| 3+ | 1.26 | (0.29-5.41) |
| P for trend | 0.776 |  |
| **Smoking History** |  |  |
| Never | 1.00 | Ref |
| Ever | 1.39 | (0.36-5.33) |
| **Cancer Type** |  |  |
| Solid | 1.00 | Ref |
| Hematological | 3.14 | (0.70-14.03) |
| **Treatment Paradigm** |  |  |
| No active treatment | 1.00 | Ref |
| Radical/Curative | 2.76 | (0.34-22.16) |
| Palliative | 3.40 | (0.50-23.20) |
| **Time since cancer** |  |  |
| ≤24 months | 1.00 | Ref |
| >24 months | 1.74 | (0.71-4.26) |
| **Performance Status** |  |  |
| 0-2 | 1.00 | Ref |
| 3+ | 0.33 | (0.04-2.64) |
| **Symptoms** |  |  |
| Cough | 1.27 | (0.53-3.01) |
| Fever | 6.21 | (1.76-21.99) |
| Dyspnoea | 2.60 | (1.00-6.76) |
| GI symptoms | 7.38 | (2.71-20.16) |
| **Time between first** |  |  |
| <7 days | 1.00 | Ref |
| 7-14 days | 0.85 | (0.28-2.54) |
| >14 days | 1.02 | (0.26-4.02) |
| **CRP (mg/L)** |  |  |
| T1 (3-41) | 1.00 | Ref |
| T2 (42-117) | 1.95 | (0.19-20.25) |
| T3 (126-508) | 9.43 | (0.73-121.12) |
| **Lymphocytes (x10<sup>9</sup>)** |  |  |
| ≤0.5 | 1.00 | Ref |
| 0.6-0.8 | 0.49 | (0.06-3.73) |
| 0.9-1.2 | 0.27 | (0.03-2.34) |
| >1.2 | 0.63 | (0.07-5.84) |
| **Albumin (g/L)** |  |  |
| T1 (20-32) | 1.00 | Ref |
| T2 (33-38) | 1.13 | (0.18-7.00) |
| T3 (39-57) | 0.07 | (0.01-0.96) |

\*Adjustment as defined by the DAG (Table 1 Appendix)

### Factors associated with COVID-19 death in cancer patients

During a median follow-up of 37 days (IQR:18-49), 34 cancer patients had died of COVID-19 (22%) (Table 5). Several cancer patient characteristics were found to be positively associated with risk of COVID-19 death: Asian ethnicity (as compared to white - HR: 3.73 (95%CI: 1.28-10.91), receiving palliative treatment (as compared to no active treatment - HR: 5.74 (95%CI: 1.15-28.79), time since cancer diagnosis >24 months (as compared to ≤24 months - HR: 2.14 (95%CI: 1.04-4.44), presenting with dyspnoea (as compared to no dyspnoea – HR: 4.94 (95%CI: 1.99-12.25), and high CRP levels (3^rd^ tertile vs 1^st^ tertile – HR: 10.35 (95%CI: 1.05-52.21)). In addition, an inverse association with death from COVID-19 was observed with levels normal albumin levels (3^rd^ tertile vs 1^st^ tertile – HR: 0.04 (95%CI: 0.01-0.042).

**Table 5:** Hazard Ratios and 95 %Confidence intervals for COVID-19 death in cancer patients.

| Variable | Number of deaths (n=34) | HR* | 95% CI |
| --- | --- | --- | --- |
| **Sex** |  |  |  |
| Male | 22 | 1.00 | Ref |
| Female | 12 | 0.73 | (0.36-1.47) |
| **Age** |  |  |  |
| ≤60 | 9 | 1.00 | Ref |
| >60 | 25 | 1.34 | (0.63-2.87) |
| **SES** |  |  |  |
| Low | 26 | 1.00 | Ref |
| Middle | 0 | N/A |  |
| High | 3 | 1.03 | (0.29-3.60) |
| **Ethnicity** |  |  |  |
| White | 21 | 1.00 | Ref |
| Black | 5 | 0.51 | (0.19-1.35) |
| Asian | 4 | 3.73 | (1.28-10.91) |
| Other | 2 | 1.52 | (0.35-6.49) |
| **Number of comorbidities** |  |  |  |
| 0 | 9 | 1.00 | Ref |
| 1 | 8 | 1.20 | (0.39-3.74) |
| 2 | 8 | 1.92 | (0.58-6.34) |
| 3+ | 9 | 1.14 | (0.34-3.82) |
| P for trend |  | 0.749 |  |
| **Smoking History** |  |  |  |
| Never | 15 | 1.00 | Ref |
| Ever | 9 | 1.00 | (0.97-1.03) |
| **Cancer Type\*\*** |  |  |  |
| Solid | 27 | 1.00 | Ref |
| Haematological | 7 | 0.22 | (0.05-1.05) |
| **Treatment Paradigm** |  |  |  |
| No active treatment | 2 | 1.00 | Ref |
| Radical/Curative | 6 | 1.35 | (0.21-8.57) |
| Palliative | 19 | 5.74 | (1.15-28.79) |
| **Time since cancer diagnosis** |  |  |  |
| ≤24 months | 15 | 1.00 | Ref |
| >24 months | 17 | 2.14 | (1.04-4.44) |
| **Performance Status** |  |  |  |
| 0-2 | 20 | 1.00 | Ref |
| 3+ | 5 | 0.56 | (0.15-2.02) |
| **Symptoms** |  |  |  |
| Cough | 16 | 1.00 | (0.48-2.09) |
| Fever | 21 | 1.63 | (0.72-3.68) |
| Dyspnoea | 21 | 4.94 | (1.99-12.25) |
| GI symptoms | 8 | 1.44 | (0.64-3.26) |
| **Time between first symptom and diagnosis** |  |  |  |
| <7 days | 23 | 1.00 | Ref |
| 7-14 days | 6 | 0.86 | (0.35-2.11) |
| >14 days | 2 | 0.54 | (0.13-2.29) |
| **CRP (mg/L)** |  |  |  |
| T1 (3-41) | 4 | 1.00 | Ref |
| T2 (42-117) | 9 | 2.87 | (0.61-13.48) |
| T3 (126-508) | 18 | 10.35 | (1.05-52.21) |
| **Lymphocytes (x10<sup>9</sup>)** |  |  |  |
| ≤0.5 | 11 | 1.00 | Ref |
| 0.6-0.8 | 12 | 0.84 | (0.21-3.38) |
| 0.9-1.2 | 4 | 0.96 | (0.20-4.57) |
| >1.2 | 4 | 0.75 | (0.14-4.12) |
| **Albumin (g/L)** |  |  |  |
| T1 (20-32) | 18 | 1.00 | Ref |
| T2 (33-38) | 8 | 0.50 | (0.17-1.47) |
| T3 (39-57) | 1 | 0.04 | (0.01-0.42) |

\*Adjustment as defined by the DAG (Table 1 Appendix)
\*\*Unadjusted due to missingness not allowing to run fully adjusted model as per the DAG.

## Discussion

Using multivariate modelling based on a directed acyclic graph, this study reports on a large cohort of COVID-19 positive cancer patients from a single institution. Low SES, hypertension and diabetes were common in cancer patients with COVID-19. Age, sex, ethnicity, SES, and current cancer treatment were found to not be associated with severity of COVID-19 infection in cancer patients. However, receipt of a cancer diagnosis more than 24 months previously (as compared to within 24 months) and presenting with fever, dyspnoea, or gastro-intestinal symptoms were linked with higher odds of developing severe illness as compared to mild/moderate COVID-19. Higher levels of CRP and ferritin were also associated with more severe COVID-19 disease in infected cancer patients. During a median follow-up of 37 days, the following cancer patient characteristics were found to be positively associated with COVID-19 death: Asian ethnicity, palliative treatment, initial cancer diagnosis >24 months, dyspnoea at presentation and high CRP levels. Normal serum albumin levels were inversely associated with death from COVID-19 in cancer patients.

### Demographic and cancer characteristics

Several retrospective cohort studies published using data from hospitals situated in Wuhan, China, Northern Italy, Canada and the USA have reported on the clinical characteristics of COVID-19 positive cancer patients with sample sizes varying from 9 to 85, two slightly larger series of >200 patients, and a big data Consortium including over 85 institutions resulting in 900 patients (5-12, 16). The median age reported in these studies was similar to our study, with a range from 63 to 72 years. A larger proportion of male patients has been observed. Lung cancer was the most commonly reported cancer in the Zhang (5), Yu (8), Yang (10) and Stroppa (11) studies, but only accounted for 11 % in our patient population. Zhang et al. (5) estimated that in their cohort 29% of patients tested positive for COVID-19 following hospital admission, whereas this was estimated at 51% in our cohort. Interpretation of this statistic is difficult given the latency between exposure and manifestations of infection, meaning patients diagnosed after admission may have been infected outside hospital. Several studies (6, 10) noted that hypertension, diabetes and coronary heart disease were the commonly reported comorbidities.

Our cancer cohort is similar in distribution of age, sex, and comorbidities to the case series reported to date. The ethnicity and SES of our COVID-19 positive cancer patients are most likely a reflection of the catchment area of our Cancer Centre in South-East London (29), covering more deprived Boroughs (Lambeth and Southwark). Based on the number of cancer patients treated at our Cancer Centre in 2019, about 49% of patients are of a White ethnic background. Variations observed with the published data in terms of cancer type, stage, and treatment may be a reflection of clinical practice (e.g. intensity of treatment and frequency of hospital visits), of relative cancer incidence, or of extent of treatment changes introduced as mitigation in the face of the emerging pandemic. For example, the most recently reported age-standardized lung cancer incidence rates for males and females in Wuhan are 54.1 and 19.1 per 100,000, whereas these are estimated to be 37.5 and 24.3 per 100,000 in London (30). Early modification and prioritization of treatment was introduced at our centre, in accordance with now-published guidance (31).

### COVID-19 characteristics and severity

Comparably to our study, both the Zhang and the Du studies also reported fever, cough, shortness of breath and dyspnea as common clinical features (5, 6). As in the Chinese cohort of 85 fatal cases, our severe COVID-19 patients had comparable laboratory findings: decreased lymphocytes, increased CRP, and decreased albumin (6).

Severe events were reported for 54% of the study population and mortality for 29% in the Zhang study, as compared to 18% and 22% in our cohort. Zhang et al. also reported that recent treatment within 14 days was associated with an increased risk of developing severe events (28 days) (5). This difference with our observations may be attributed to different definitions of severe events, as it was not entirely clear how these were defined by Zhang et al. As highlighted by Wynants et al in their assessment of current statistical models published for COVID-19 (19), there is a need for consistent use of outcome definitions. However, our observations of a positive association with CRP levels is in line with most COVID-19 studies published to date (32). Apart from the CCC-19 Consortium (16), no study to date has specifically looked at COVID-19 severity at presentation in COVID-19 positive cancer patients and hence our observation of an association with time since cancer diagnosis and presenting symptoms needs further validation in other large cohorts with homogenous definitions of inclusion criteria, testing strategies, and outcomes measures. However, it is possible that time since cancer diagnosis is also a reflection of extent of disease and progression along the palliative patient pathway from diagnosis to death.

### COVID-19 death

The study by Yu et al. reported three deaths (25%) (8). In the larger series from New York, Mehta et al reported an overall case fatality rate of 28%, with 37% for haematological malignancies and 25% for solid tumours (14). The CCC-19 Consortium reported a 30-day mortality rate due to COVID-19 of 13% (16). In our cohort the overall case fatality rate was 22%, with 25% for haematological cancers and 21% for solid tumours. As more than 85 institutions were included in the CCC-19 Consortium (16), it is possible that differences in COVID-19 practice as well as cancer treatments between the numerous centres may explain the slightly lower death rate as compared to reports of single centre study. Moreover, our median follow-up is 37 days as compared to 21 days for the Consortium. The heterogeneity between centres may also explain why performance status was found to be associated with COVID-19 outcomes, an observation not identified in our single centre cohort.

Our observations of Asian ethnicity being associated with increased mortality from COVID-19 in cancer patients is of interest, given the recent speculations about the disproportionate effects of COVID-19 on Black, Asian and minority ethnic communities (33) as well as the confounding factor of vitamin D deficiency (34). However, longer follow-up studies are required to disentangle the association between ethnicity and COVID-19 death in cancer patients

### Strengths and limitations

Whilst this is one of the largest single centre COVID-19 positive cancer cohorts to date, our sample size is still relatively modest and hence confidence intervals for some statistically significant observations are still wide. No firm conclusions in terms of prognostic modelling can be drawn as of yet (19). Current analyses were aimed at hypothesis generation about patient or tumour characteristics indicative of severity of or death from COVID-19 in the cancer context. Our data for some of the patient characteristics is limited, for example smoking status was missing for 29% of patients and hence likely underestimates the proportion of ever smokers. COVID testing in the UK has only been implemented gradually during the period of our data collection, and there is selection bias in favour of patients being tested as inpatients. Our analysis is likely to have missed cancer outpatients under our care diagnosed with COVID-19 at other hospitals – however this is most likely to be an even more important issue for global Consortia with many hospitals only adding a few cases to the overall dataset. It is a strength of our study that we used clearly defined definitions of COVID-19 severity, as well as a DAG to develop the different models, as to date very limited knowledge is available regarding the intersection between COVID-19 and cancer (19). Detailed information on our modelling will help comparison with future studies with larger sample sizes and longer follow-up.

## Conclusion

Our analysis of one of the largest single centre series of COVID-19 positive cancer patients to date confirms a similar distribution of age, sex, and comorbidities as reported for other populations. Reflecting the general population, presenting with fever, dyspnoea, or gastro-intestinal symptoms, higher levels of CRP or ferritin were also indicators of COVID-19 severity in the cancer population. Similarly, we noted that dyspnoea at presentation, high CRP levels, and low levels of albumin were associated with death from COVID-19. With respect to cancer specific observations, patients who have lived longer with their cancer were found to be more susceptible to a greater infection severity, possibly reflecting the effect of more advanced malignant disease, as almost half of the severe cohort were on third line metastatic treatment, or the impact of this infection. The latter was also found to be associated with COVID-19 death in cancer patients, as were Asian ethnicity and palliative treatment. Further validation will be provided from other large case series, as well as from those including longer follow-up, to provide more definite guidance for oncological care.

## Data Availability

Data can be requested by contacting

## Acknowledgements

The research was supported by the National Institute for Health Research (NIHR) Biomedical Research Centre (BRC) based at Guy’s and St Thomas’ NHS Foundation Trust and King’s College London (IS-BRC-1215-20006). The authors are solely responsible for study design, data collection, analysis, decision to publish, and preparation of the manuscript. The views expressed are those of the authors and not necessarily those of the NHS, the NIHR or the Department of Health. We also acknowledge support from Cancer Research UK King’s Health Partners Centre at King’s College London and Guy’s and St Thomas’ NHS Foundation Trust Charity Cancer Fund.

We are grateful to Graham Roberts for providing us with the descriptive statistics of the cancer patients treated at our Cancer Centre.

## Author contribution

Data collection: BR, CM, PR, DC, HW, FC, AH, KZ, FR, SLA, JD, DS, ML, SG, ES, AD, SG, DE, PF, DW, KR, MG, KB, RD, MS, AS,

Study design: BR, CM, SP, IS, PR, JS, SD, MVH

Data analysis: BR, CM, MVH, SD

Manuscript drafting: MVH, BR, CM, SD, SP, RS, PR, JS, SK, CH

Final approval of manuscript: All authors

## Ethics approval

Guy’s Cancer Cohort, a research ethics committee approved research database (Reference number: 18/NW/0297) of all routinely collected clinical data of cancer patients at Guy’s and St Thomas’ NHS Foundation Trust (GSTT), forms the basis of this observational study.

## Data availability

Data can be obtained by researchers via an application to the Access Committee of Guy’s Cancer Cohort. An application form can be obtained via

